# Prognostic Implications of Codon-Specific *KRAS* Mutations in Localized and Advanced Stages of Pancreatic Cancer

**DOI:** 10.1101/2025.02.03.25321601

**Authors:** Sara Raji, Hamed Zaribafzadeh, Tyler Jones, Elishama Kanu, Kunling Tong, Ashley Fletcher, T. Clark Howell, Shannon J. McCall, Jeffrey R. Marks, Bruce Rogers, Donna Niedzwiecki, Peter J. Allen, Daniel P. Nussbaum, Zahra Kabiri

**Affiliations:** Department of Surgery, Duke University School of Medicine, Durham, NC, USA; Department of Biostatistics and Bioinformatics, Duke University School of Medicine, Durham, NC, USA; Department of Pathology, Duke University School of Medicine, Durham, NC, USA

## Abstract

**Introduction:** While *KRAS* mutations represent the primary oncogenic driver in pancreatic ductal adenocarcinoma (PDAC), the association between codon-specific alterations and patient outcomes remains poorly elucidated, largely due to a lack of datasets coupling genomic profiling with rich clinical annotations across disease stages.

**Patients and Methods:** We utilized AACR’s GENIE Biopharma Consortium Pancreas v1.2 dataset to test the associating of codon-specific *KRAS* mutations with clinicogenomic features and patient outcomes in PDAC patients diagnosed with localized (stages I-III) and advanced disease (stage IV). Overall survival was compared using Kaplan–Meier and multivariable Cox proportional hazards methods.

**Results:** Among 1,032 eligible patients, 949 (92%) exhibited mutant *KRAS*. These mutations were predominantly observed at G12D (n=390, 41%), G12V (n=305, 32%), and G12R (n=149, 16%). In the group of patients who presented with localized disease, those with G12V mutation had notably longer survival compared to G12D mutation (*P* = 0.002). In contrast, patients with G12V mutation who presented with metastatic disease experienced shorter overall survival compared to those with G12R (*P* = 0.005), and G12D mutations (*P* = 0.009). Furthermore, no significant differences were observed in the frequencies of co-altered driver genes, including *TP53*, *CDKN2A*, and *SMAD4*, across the different *KRAS* mutations.

**Conclusions:** These findings demonstrated that codon-specific *KRAS* mutations impact PDAC outcomes differently based on disease stage at diagnosis. As studies testing *KRAS* inhibitors continue to emerge and mature, these data offer important contextual insights regarding survival outcomes associated with codon-specific *KRAS* mutations based on existing therapeutic approaches.

## Introduction

Pancreatic ductal adenocarcinoma (PDAC) has a five-year survival rate of approximately 10%, mainly due to the lack of effective methods for early detection and treatment (1,2). Over 80% of patients present with advanced disease at the time of diagnosis, and outside of clinical trials, standard treatment is cytotoxic chemotherapy that provides limited benefit (3,4). In patients diagnosed with localized disease, the combination of surgical resection and systemic chemotherapy can improve survival in select patients, however, the vast majority of patients treated with curative intent develop disease recurrence (5). These dismal outcomes highlight the need for improved understanding of the biologic underpinnings of PDAC.

*KRAS* is mutated in over 90% of all PDAC patients, serving as the primary oncogenic driver in this cancer. These *KRAS* mutations predominantly occur at codon 12, with less frequent mutations at other codons. By far the most common codon-specific mutations are G12D (∼40%), G12V (∼30%), G12R (∼15%), and Q61 (H, K, L, R) (∼6%) (6,7). In preclinical studies, the various codon-specific *KRAS* mutations have demonstrated unique biochemical properties, suggesting that they may similarly promote differential clinicopathological features in patients (8,9). Decades of research on the prognostic role of specific *KRAS* mutations in various cancers have significantly deepened our understanding of their importance in predicting clinical outcomes (10–13). However, results remain conflicting regarding their prognostic role in pancreatic cancer, largely due to a lack of large datasets that couple genomic profiling with rich clinical annotations (6,7,14–19). Additionally, previous studies have typically focused on either early or late stages of the disease, rather than across all stages.

The American Association for Cancer Research’s Project GENIE (Genomics Evidence Neoplasia Information Exchange) is a publicly accessible cancer registry that includes robust clinicogenomic data derived from multiple leading international cancer centers. In this study, we conducted a retrospective analysis to elucidate the impact of codon-specific mutations on PDAC patient outcomes across disease stages, leveraging the detailed demographic, clinical, and treatment-level annotations that have not previously been available in other genomic profiling efforts.

## Methods and Materials

### Patient selection from AACR Project GENIE Database

We used the Pancreas v1.2-consortium multi-institutional and multilayer database from the AACR’s GENIE Biopharma Consortium (BPC) (20). In summary, the GENIE BPC PANC cohort comprises 1130 samples obtained from 1109 patients meeting specific eligibility criteria as previously reported (21,22). Samples with cancer types other than pancreatic adenocarcinoma, missing *KRAS* mutation data, those with the co-occurrence of *KRAS* mutations in the different category (G12D, G12V, G12R, Q61, other, and wild-type) were excluded. The research ethics committees of all institutions involved granted approval for the GENIE BPC project.

### Clinical data

The clinical data was carefully curated using PRISSMM (Pathology; Radiology; Imaging; Signs and Symptoms; tumor Markers; Medical oncology assessments) framework, which is a system designed for extracting clinical data from longitudinal electronic health records (EHR) and has been previously described (23). Demographic characteristics, staging, and oncology-related outcomes were evaluated. Patients were categorized into cohorts based on stage at diagnosis: localized (stages I-III) and metastatic (stage IV). Patients were also evaluated for systematic therapies, which were categorized as follows: neoadjuvant chemotherapy or adjuvant chemotherapy (localized disease), and first-line treatment (metastatic disease).

### Genomic data

We extracted publicly available mutation data in the mutation annotation or variant cell format from four centers through Synapse using the cBioPortal interface. Comprehensive details regarding the clinical sequencing pipelines of each center can be found in the GENIE data guide (GENIE 15.1 public release; https://www.aacr.org/wp-content/uploads/2024/02/15.0-public_data_guide-.pdf). This cohort included genomic data on 39 genes that were assessed in all oncopanels across the four centers. Copy number variation (CNVs) data for these genes were available for 975 patients. The term “altered” was used for the tumor suppressor genes *CDKN2A*, *SMAD4*, and *TP53* if any copy number loss or mutations were observed. We calculated the genomic alterations based on changes in copy number variations or mutations in 35 non-driver genes, excluding *KRAS*, *CDKN2A*, *SMAD4*, and *TP53*. **Supplementary Table S1** lists all gene sequencing panels used by the four centers.

### Statistical analyses

Descriptive statistics, including median/quartile and percentage were used to summarize baseline patient and clinical characteristics. Comparisons between groups were performed using Chi-square or Fisher’s exact tests for categorical variables and the Wilcoxon rank-sum test for nonparametric continuous variables. False Discovery Rate (FDR) correction was applied for multiple comparisons. Survival probabilities were estimated using the Kaplan-Meier method, and differences in survival were compared using the log-rank test. Overall survival (OS) was defined as the time from initial diagnosis to either the date of death or last known follow-up, and patients alive at the last follow-up were considered censored in the analysis. The Cox proportional hazards model was used to simultaneously assess prognostic factors and estimate hazard ratios across codon-specific mutations while controlling for other clinical features.

All statistical analyses were two-sided, with a p-value threshold of 0.05 considered statistically significant. Analyses were performed using IBM SPSS software, version 28.0.1.1, GraphPad Prism, version 10.2.2. and Python.

### Data availability

The data are publicly available through cBioPortal (https://genie.cbioportal.org). The clinical and genomic data can be obtained from Synapse (https://www.synapse.org/genie) upon creating a synapse account.

## Results

### Patient characteristics

A total of 1032 patients were included in this study, consisting of 464 females (45%) and 568 males (55%), with a median age of 65 (IQR: 57-72). Most of the patients were non-Hispanic white (837, 81%). Patients data were derived from four centers: Memorial Sloan Kettering (MSK, 500, 48%), Dana Farber Cancer Institute (DFCI, 421, 40%), Vanderbilt Ingram Cancer Center (VICC, 63, 6.1%), and University Health Network (UNH, 57, 5.5%). Patients were relatively evenly split between those diagnosed with localized disease (stage I-III, 565, 55%) and those diagnosed with metastatic disease (stage IV, 467, 45%). Complete baseline patient characteristics stratified by stage at diagnosis are listed in **Table 1**. *KRAS* mutations were present in 949 patients (92%) and absent in 83 patients (8%). Among the *KRAS* mutations, the most common was G12D, found in 390 cases (37.8%), followed by G12V in 305 cases (29.6%), G12R in 149 cases (14.4%), Q61 (H, R, K, and L) in 74 cases (7.2%), and other less frequent mutations in 31 cases (3%). A detailed list of *KRAS* mutations is provided in **Supplementary Table S2**. Complete baseline patient characteristics according to codon-specific *KRAS* mutations are shown in **Table 2**.

**Table 1.** GENIE cohort patient characteristics.

| Characteristic | Overall<br>(N=1,032), N (%) | Stages I-III (N=565),<br>N (%) | Stage IV<br>(N=467),<br>N (%) |
| --- | --- | --- | --- |
| **Age (Q1-Q3)** | 65 (57-72) | 66 (58-72) | 64 (57-71) |
| **Sex** |  |  |  |
| Female | 464 (45%) | 263 (47%) | 201 (43%) |
| Male | 568 (55%) | 302 (53%) | 266 (57%) |
| **Race/Ethnicity** |  |  |  |
| Asian | 37 (3.6%) | 16 (2.8%) | 21 (4.5%) |
| Hispanic/Latinx | 85 (8.2%) | 48 (8.5%) | 37 (7.9%) |
| Non-Hispanic Black | 36 (3.5%) | 21 (3.7%) | 15 (3.2%) |
| Non-Hispanic White | 837 (81%) | 463 (82%) | 374 (80%) |
| Other | 37 (3.6%) | 17 (3%) | 20 (4.3%) |
| **Center** |  |  |  |
| DFCI | 412 (40%) | 212 (38%) | 200 (43%) |
| MSK | 500 (48%) | 287 (51%) | 213 (46%) |
| UHN | 57 (5.5%) | 32 (5.7%) | 25 (5.4%) |
| VICC | 63 (6.1%) | 34 (6.0%) | 29 (6.2%) |
| **KRAS status** |  |  |  |
| Wild-type | 83 (8%) | 44 (7.8%) | 39 (8.4%) |
| G12D | 390 (37.8%) | 214 (37.9%) | 176 (37.7%) |
| G12V | 305 (29.6%) | 172 (30.4%) | 133 (28.5%) |
| G12R | 149 (14.4%) | 85 (15%) | 64 (13.7%) |
| Q61 | 74 (7.2%) | 32 (5.7%) | 42 (9%) |
| Other | 31 (3%) | 19 (3.4%) | 14 (3%) |
| **PDAC driver genes** |  |  |  |
| **TP53** |  |  |  |
| Altered | 759 (73.5%) | 402 (71.2%) | 357 (76.4%) |
| Unknown | 57 (5.5%) | 32 (5.7%) | 25 (5.4%) |
| **CDKN2A** |  |  |  |
| Altered | 402 (39%) | 172 (30.4%) | 230 (49.2%) |
| Unknown | 57(5.5%) | 32 (5.7%) | 25 (5.4%) |
| **SMAD4** |  |  |  |
| Altered | 379 (36.7%) | 183 (32.4%) | 196 (42%) |
| Unknown | 57(5.5%) | 32 (5.7%) | 25 (5.4%) |

Abbreviations: DFCI, Dana Farber Cancer Institute; MSKCC, Memorial Sloan Kettering Cancer Center; UHN, Princess Margaret Cancer Centre-University Health Network; VICC, Vanderbilt Ingram Cancer Center.

**Table 2.** Baseline patient characteristics according to codon-specific *KRAS* mutations.

| Variable | G12D, N=390 (%) | G12V, N=305 (%) | G12R, N=149 (%) | Q61, N=74 (%) | Other, N=32 (%) | WT, N=83 (%) | * *P* value |
| --- | --- | --- | --- | --- | --- | --- | --- |
| **Age** |  |  |  |  |  |  |  |
| <65 | 210 (53.8) | 133 (43.6) | 61 (40.9) | 26 (35.1) | 11 (35.5) | 54 (65.1) | **<0.001** |
| ≥65 | 180 (46.2) | 172 (56.4) | 88 (59.1) | 48 (64.9) | 20 (64.5) | 29 (34.9) |  |
| **Sex** |  |  |  |  |  |  |  |
| Male | 204 (52.3) | 183 (60) | 72 (48.3) | 37 (50) | 18 (58.1) | 54 (65.1) | **0.048** |
| Female | 186 (47.7) | 122 (40) | 77 (51.7) | 37 (50) | 13 (41.9) | 29 (34.9) |  |
| **Race** |  |  |  |  |  |  |  |
| White | 330 (84.6) | 245 (80.3) | 118 (79.2) | 63 (85.1) | 24 (77.4) | 57 (68.7) | **0.023** |
| Other | 60 (15.4) | 60 (19.7) | 31 (20.8) | 11 (14.9) | 7 (22.6) | 26 (31.3) |  |
| **Center** |  |  |  |  |  |  |  |
| DFCI | 164 (42.1) | 113 (37.0) | 59 (39.6) | 30 (40.5) | 14 (45.2) | 32 (38.6) | **0.002** |
| MSK | 190 (48.7) | 158 (51.8) | 72 (48.3) | 37 (50.0) | 11 (35.5) | 32 (38.6) |  |
| UHN | 12 (3.1) | 16 (5.2) | 13 (8.7) | 2 (2.7) | 1 (3.2) | 13 (15.7) |  |
| VICC | 24 (6.2) | 18 (5.9) | 5 (3.4) | 5 (6.8) | 5 (16.1) | 6 (7.2) |  |
| **Stage at diagnosis** |  |  |  |  |  |  |  |
| I-III | 214 (54.9) | 172 (56.4) | 85 (57) | 32 (43.2) | 18 (58.1) | 44 (53) | 0.4 |
| IV | 176 (45.1) | 133 (43.6) | 64 (43) | 42 (56.8) | 13 (41.9) | 39 (47) |  |
| **Metastasis** |  |  |  |  |  |  |  |
| Liver | 276 (70.8) | 216 (70.8) | 88 (59.1) | 53 (71.6) | 18 (58.1) | 59 (71.1) | 0.07 |
| Thorax | 160 (41) | 127 (41.6) | 57 (38.3) | 27 (36.5) | 11 (35.5) | 41 (49.4) | 0.5 |

Abbreviations: DFCI, Dana Farber Cancer Institute; MSKCC, Memorial Sloan Kettering Cancer Center; UHN, Princess Margaret Cancer Centre-University Health Network; VICC, Vanderbilt Ingram Cancer Center.

### Genomic landscape of PDAC in the GENIE dataset

To investigate the genomic alterations in this cohort, we focused on a panel of 39 genes that had complete somatic mutation and CNV data across all four centers. Other than *KRAS*, the most commonly altered genes were: *TP53* in 759 (73.5%), *CDKN2A* in 402 (39%), and *SMAD4* in 379 (36.7%). The genomic landscape, stratified by stage at diagnosis and codon-specific *KRAS* mutations, are summarized in **Fig. 1**.

**Fig. 1.**
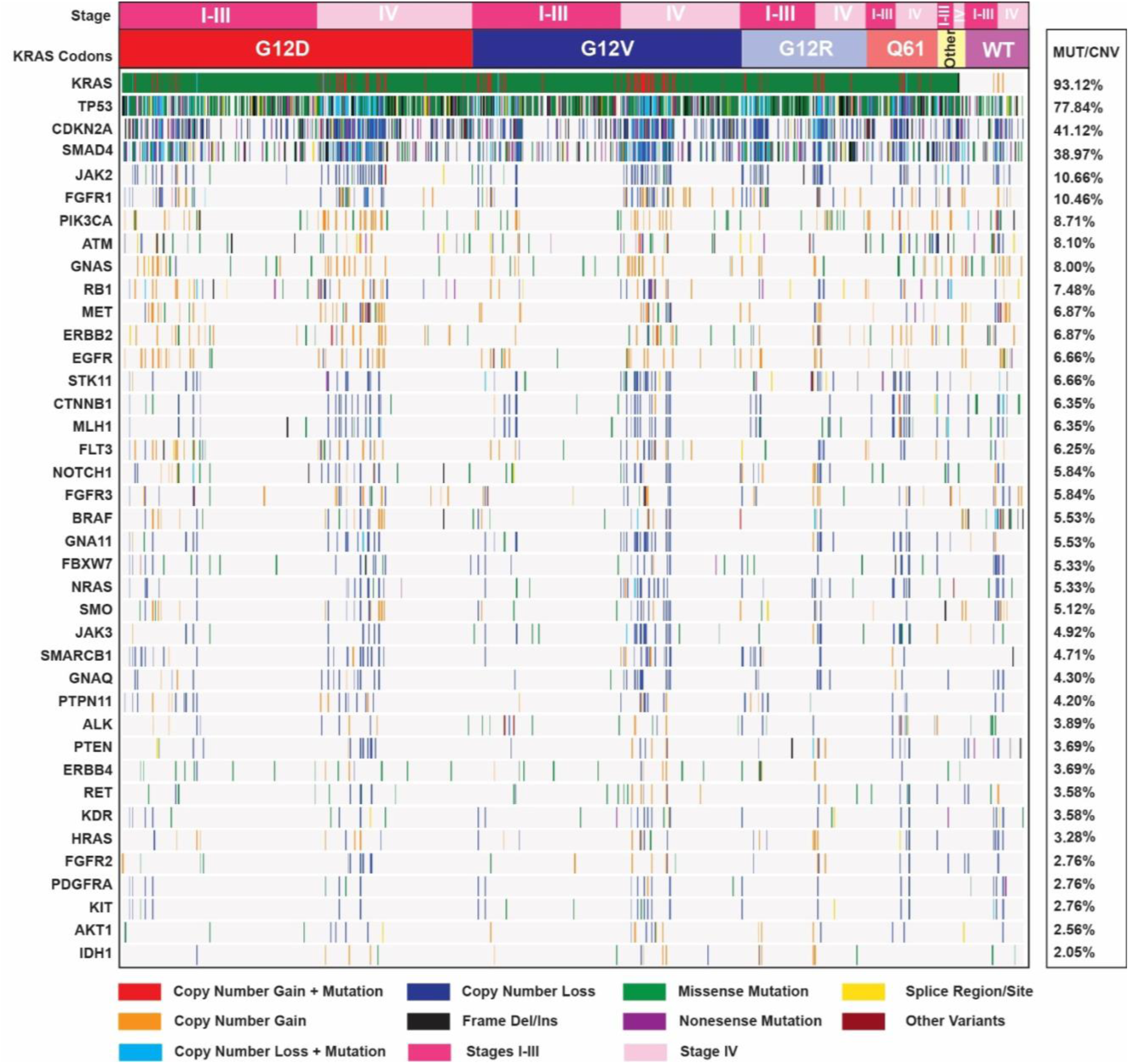
Genomic landscape of GENIE BPC PANC cohort. The oncoplot illustrates genomic alterations in 39 genes in the GENIE dataset per patient for 975 patients. The distribution of codon-specific *KRAS* mutations across stages I-III and stage IV PDAC patients are shown at the top. The right panel displays the frequency of copy number variations or mutations in the cohort. The specific types of mutations as well as copy number loss and gain in the oncoplot are listed at the bottom.

Given that PDAC tumors often exhibit co-occurring alterations in these driver genes, we next assessed the frequency of genomic alterations in *TP53, CDKN2A*, and *SMAD4* in the context of wild-type and mutant *KRAS* **(Fig. 2A and 2B)**, as well as the most common codon-specific *KRAS* mutations **(Fig 2C-F)**. Compared to patients with wild-type *KRAS*, patients with *KRAS* mutations were more likely to harbor co-occurring alterations in *TP53* (80.3% vs. 45.7%; *P* < 0.001), *TP53* and *CDKN2A* (36.6% vs. 14.2%; *P* < 0.001), *TP53* and *SMAD4* (33% vs. 12.8%; *P* = 0.003), as well as alterations in *TP53*, *SMAD4*, and *CDKN2A (*20% vs. 7.1%; *P* = 0.004). The absence of any of these co-alterations was less common in patients with mutant *KRAS* compared to wild-type *KRAS* (10.4% vs. 27.4%; *P* = 0.003). Notably, no significant differences in co-alterations were observed between the codon-specific *KRAS* mutations (all *P* > 0.05).

**Fig. 2.**
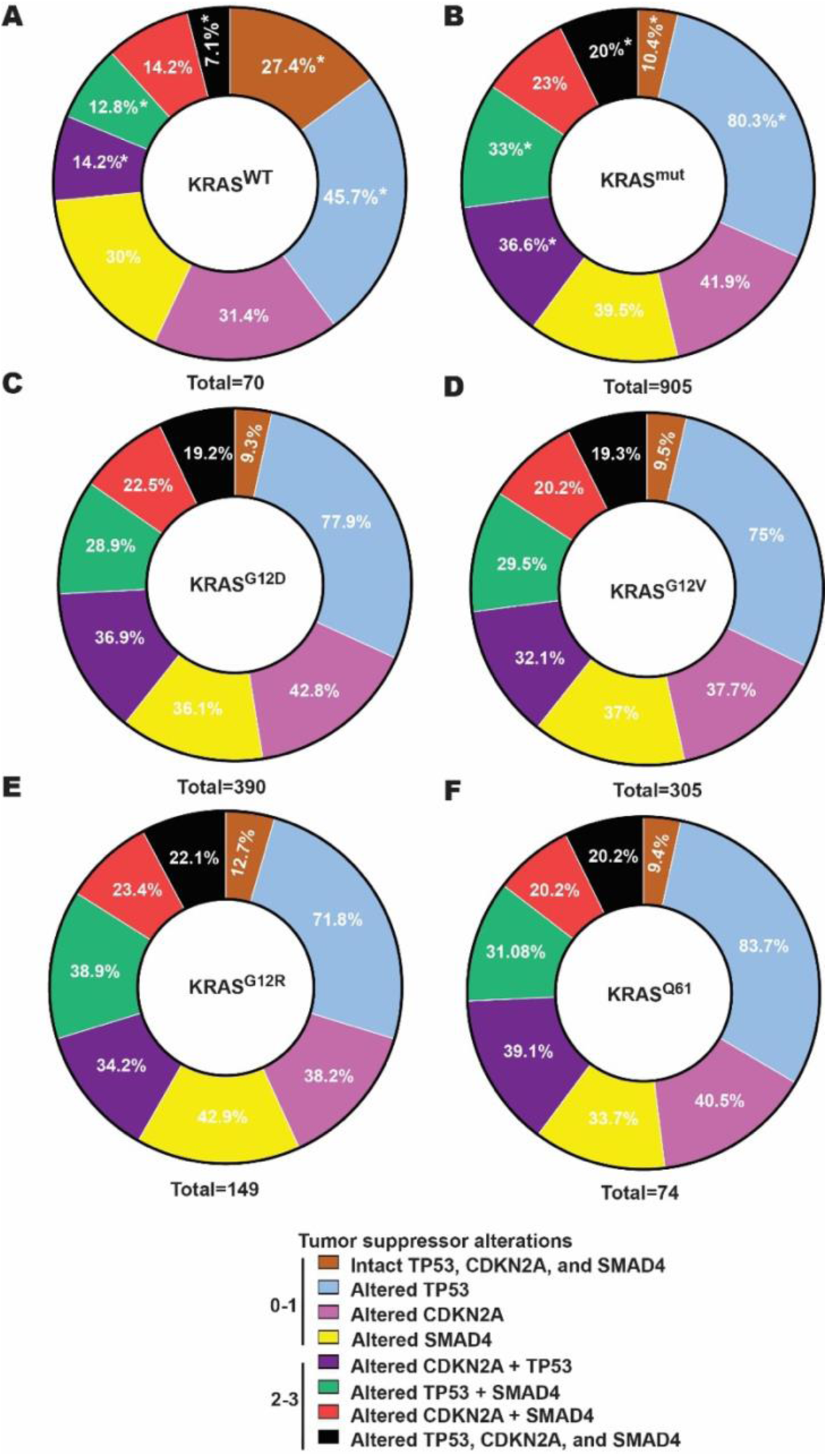
Frequencies of tumor suppressor alterations. Combination of altered *TP53*, *CDKN2A*, and *SMAD4* in the presence of mutated, wild-type and different *KRAS* codon mutations tumors. Donut charts representing combination of *TP53*, *CDKN2A,* and *SMAD4* mutation for each of **(A)** wild-type *KRAS*, **(B)** *KRAS* mutant, **(C)** *KRAS* G12D, **(D)** *KRAS* G12V, **(E)** *KRAS* G12R, and **(F)** *KRAS* Q61. Chi Square test was performed between wild-type and *KRAS* mutant groups to compare tumor suppressor alterations. *, P value < 0.05. Chi squares between different *KRAS* codon mutations revealed no significant differences for tumor suppressor alterations. WT, wild-type; mut, mutant.

We then investigated the impact of alterations in *TP53*, *CDKN2A*, and *SMAD4* on PDAC outcomes, independent of *KRAS* mutation status. This revealed that each of these alterations had a negative prognostic impact on survival. Specifically, patients with altered compared to wild-type *TP53* had a median survival of 19.4 months versus 27.8 months, respectively (HR 1.44; 95% CI 1.23-1.69; *P* = 0.001). Similarly, patient with altered compared to wild-type *CDKN2A* (n=402) had a median survival of 15.7 months versus 25.3 months, respectively (HR 1.70; 95% CI 1.46-1.98; *P* = 0.001), and patients with altered compared to wild-type *SMAD4* had a median survival of 19.2 months compared to 22.5 months, respectively (HR 1.22; 95% CI 1.05-1.41; *P* = 0.006) **(Supplementary Fig. S2A-D).** In patients with *KRAS*-mutant tumors, we observed that the absence of alteration in *TP53*, *CDKN2A*, and *SMAD4* was associated with the best survival (28.3 months), while patients with tumors containing just one of these altered tumor suppressors had significantly reduced overall survival. Notably, the worst survival outcomes were observed in patients with tumors that harbored altered *CDKN2A*, with similar survival whether CDKN2A was altered alone (15.55 months), concurrently with *SMAD4* (14.4 months), or concurrently with *TP53* (14 months) **(Supplementary Fig. S2E and Table S3)**. Thus, in patients with *KRAS*-mutant PDAC, loss of *CDKN2A* appears to confer a particularly dismal prognosis.

### Association of Codon-specific *KRAS* mutations with overall survival in localized and advanced disease stages

Next, we investigated whether different *KRAS* mutations have distinct impacts on overall survival. In the main cohort (not stratified by stage), there were no statistically significant differences in overall survival based on codon-specific *KRAS* mutations (log-rank *P* = 0.2) **(Fig. 3A and 3B)**, nor were the frequencies of co-alterations in the common tumor suppressor genes significantly different among the codon-specific *KRAS* mutation groups **(Fig. 3C and Supplementary Table S4)**. Interestingly, however, codon-specific *KRAS* mutations were associated with distinct survival outcomes in patients diagnosed with localized **(Fig. 3D and 3E)** compared to metastatic disease (**Fig. 3G and 3H)**, suggesting a prognostic role for these mutations based on disease stage.

**Fig. 3.**
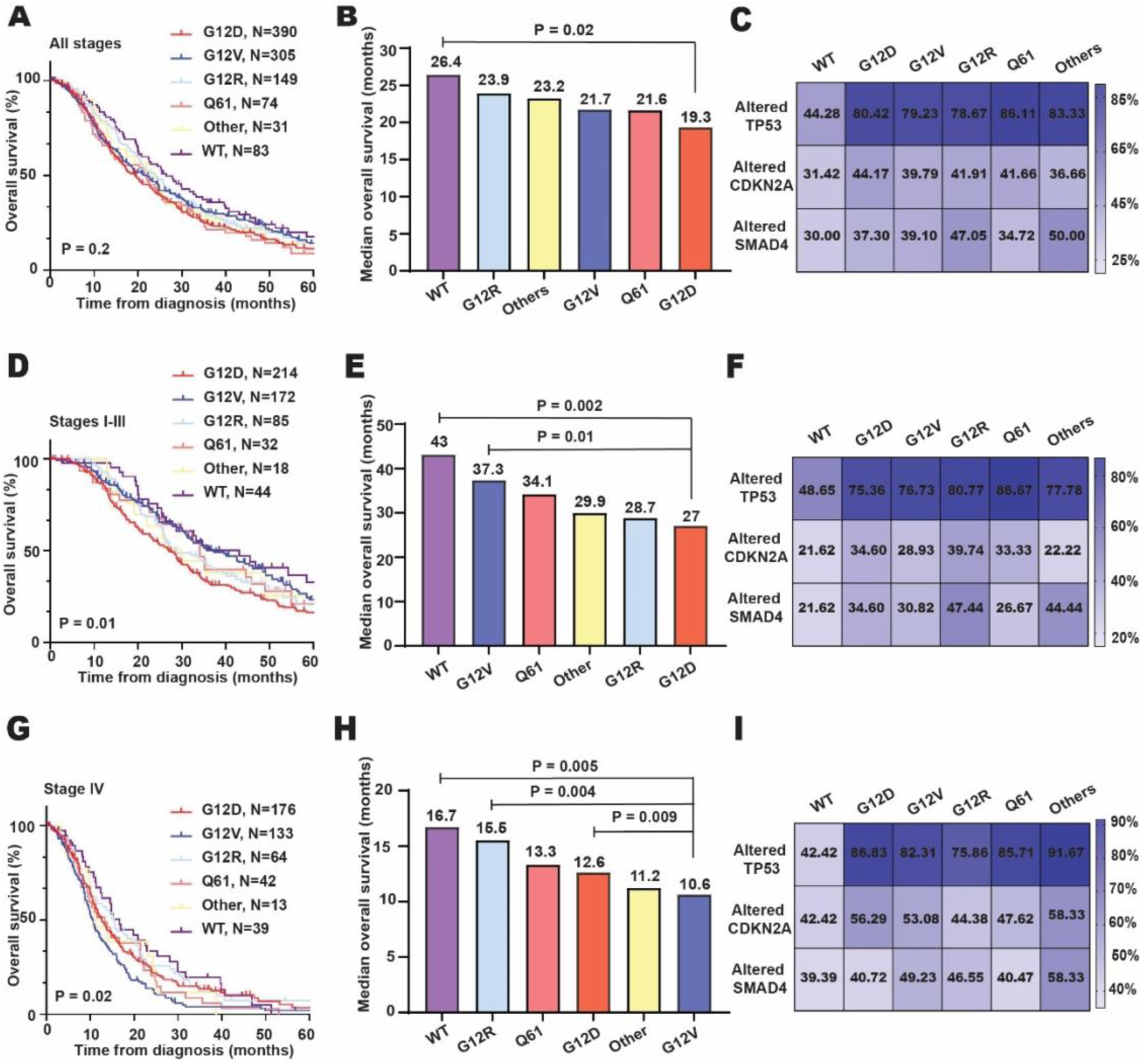
Association of *KRAS* mutations with overall survival. Overall survival analyses comparing all codon-specific *KRAS* mutations in (**A**) all stages, (**D**) stages I-III, and (**G**) stage IV of PDAC patients. Bar charts show the median overall survival of different codon-specific *KRAS* mutations and *KRAS WT* (wild-type) in (**B**) all stages, (**E**) stages I-III, and (**H**) stage IV of PDAC patients. Survival was analyzed using Kaplan Meier method and compared between groups using log-rank test. Heatmaps describe the frequency distribution of the codon-specific *KRAS* mutations (G12D, G12V, G12R, Q61, and other) and wild-type *KRAS* among PDAC patients of all stages **(C)**, stages I-III **(F)**, and stage IV **(I)** with commonly co-altered tumor suppressor genes (*TP53*, *CDKN2A*, and *SMAD4*). No adjustment was made for multiple comparisons. The detailed information is provided in **Supplementary Table S4**.

In patients diagnosed with stage I-III disease, the *KRAS* G12V mutation was associated with significantly longer survival compared to the G12D mutation (37.3 months vs. 27 months; HR 0.69; 95% CI 0.55-0.87; *P* = 0.001) (**Fig. 3E**). We investigated whether this difference in survival could be explained by differential frequencies of *TP53*, *CDKN2A*, and *SMAD4* co-alterations among codon-specific *KRAS* mutations. However, the frequencies of co-alterations in these tumor suppressor genes were not significantly different among the codon-specific *KRAS* mutation groups (**Fig. 3F and Supplementary Table S5**).

Conversely, in patients diagnosed with stage IV disease, the *KRAS* G12V mutation was associated with shorter survival (10.6 months) compared to G12D (12.6 months; HR 1.37; 95% CI 1.07-1.75; *P* = 0.009) and G12R (15.5 months; HR 1.54; 95% CI 1.13-2.10; *P* = 0.005) mutations **(Fig. 3H).** Similar to all stages and stages I-III, no significant differences were observed for the frequencies of tumor suppresser alternations among codon-specific *KRAS* mutations **(Fig. 3I and Supplementary Table S6)**.

### Paradoxical impact of *KRAS* G12V versus G12D mutation on survival based on stage at diagnosis

*KRAS* G12D and G12V mutations are the most frequent *KRAS* mutations observed in PDAC, occurring in over 70% of cases. Our survival analyses revealed a paradoxical impact of *KRAS* G12V compared to G12D mutations in patients diagnosed with localized versus metastatic disease. We aimed to determine whether this observation was independently associated with *KRAS* codon status or influenced by other factors, such as genomic co-alterations in *TP53*, *CDKN2A*, and *SMAD4* genes, patterns of distant metastasis, or treatment regimens. To test the independent associations between *KRAS* G12V and G12D status and survival outcomes, we applied univariable and multivariable Cox regression models **(Fig. 4 and Supplementary Fig. S3)**.

**Fig. 4.**
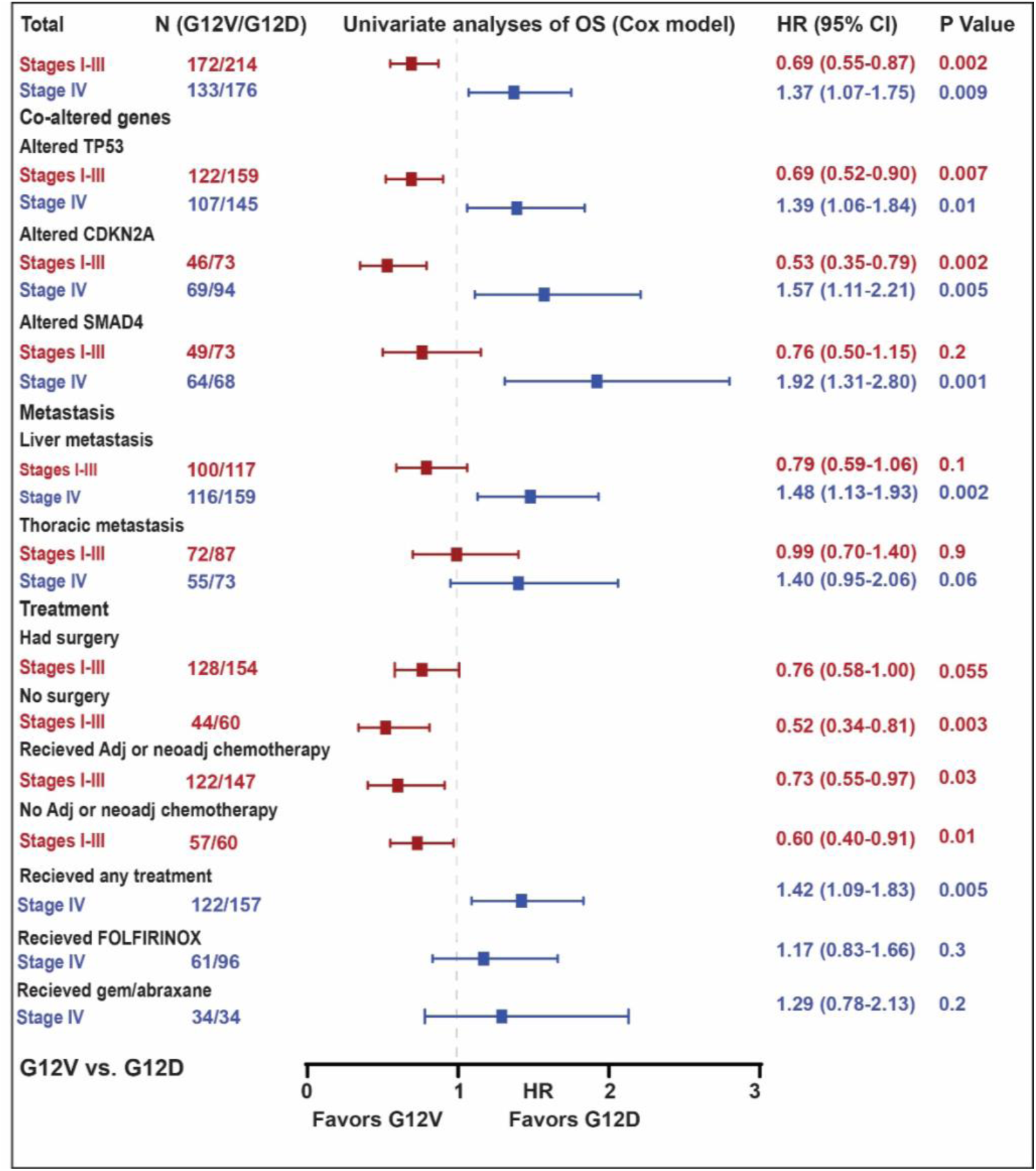
Forest plot of univariable Cox proportional hazards models of overall survival within subgroups. Univariable analyses of overall survival in *KRAS* G12V versus G12D status within different stages, alterations in *TP53*, *CDKN2A*, and *SMAD4*, liver and thoracic metastasis, surgical status, adjuvant or neoadjuvant chemotherapy status in stages I-III, and any first-line treatment, first-line FOLFIRINOX status, and first-line gem/abraxane status in stage IV disease. Survival analyses were done using Cox proportional hazards model and *P* values were calculated by Wald test, overall survival; HR, hazard ratio; CI, confidence interval; Adj, adjuvant; neoadj, neoadjuvant; gem/abraxane, gemicitabibe/abraxane.

In univariable analysis of patients diagnosed with stage I-III disease, the *KRAS* G12V mutation was associated with longer overall survival compared to *KRAS* G12D (HR 0.69; 95% CI 0.55-0.87; *P* = 0.002). This association was conserved in the *TP53* altered subgroup (HR 0.69; 95% CI 0.52-0.90; *P* = 0.007) and the *CDKN2A* altered subgroup (HR 0.53; 95% CI 0.35-0.79; *P* = 0.002), as well as across different treatment regimens. In patients with *SMAD4* co-alterations, those who eventually developed liver and thoracic metastasis, and in patients who underwent surgery, the G12V mutation was again associated with improved overall survival, but these differences did not reach statistical significance: 0.76 (95% CI 0.50-1.15, *P* = 0.2), 0.79 (95% CI 0.59-1.06, *P* = 0.1), 0.99 (95% CI 0.70-1.40, *P* = 0.9), and 0.76 (95% CI 0.58-1.00, *P* = 0.055), respectively.

Conversely, in univariable analysis of patients diagnosed with metastatic disease, the *KRAS* G12V mutation was associated with worse survival compared to the G12D mutation (HR 1.37; 95% CI 1.07-1.75; *P* = 0.009). This trend remained consistent in the presence of alterations in *TP53* (HR 1.39; 95% CI 1.06-1.84; *P* = 0.01), *CDKN2A* (HR 1.57; 95% CI 1.11-2.21; *P* = 0.005), and *SMAD4* (HR 1.92; 95% CI 1.31-2.80; *P* = 0.001), as well as in patients diagnosed with liver metastases (HR 1.48; 95% CI 1.13-1.93; *P* = 0.002) and patients that received any first-line systemic therapy (HR 1.42; 95% CI 1.09-1.83; *P* = 0.005). No statistically significant difference in overall survival between *KRAS* G12D and G12V mutations were detected within stage IV groups that received specific systemic regimens, possibly due to the limited size of each of these cohorts.

These findings were further confirmed with multivariable analysis using Cox proportional hazards models adjusted for age at diagnosis, sex, race, and treatment center **(Supplementary Fig. S3)**. The treatment profiles, including adjuvant and neoadjuvant chemotherapies for stages I-III and first-line chemotherapies for stage IV among patients with *KRAS* G12D or G12V mutations are detailed in **Supplementary Table S7, S8, and S9**, respectively.

### The paradoxical impact of *KRAS* G12V mutation on survival is associated with genomic alterations in non-driver genes

Considering the paradoxical impact of *KRAS* G12V mutation status on PDAC prognosis based on stage at diagnosis, we investigated whether the frequency of CNVs or mutations (referred here as genomic alterations) in the 35 non-driver genes evaluated across all gene panels correlated with different clinical outcomes. Interestingly, the frequency of genomic alterations in 15 out of the 35 genes was significantly lower in G12V patients diagnosed with stage I-III disease (those with the best clinical outcomes) compared to patients diagnosed with stage IV disease (those with worst clinical outcomes) **(Fig. 5A, and Supplementary Table S10).** However, no significant differences in genomic alterations of non-driver genes were observed between patients with *KRAS* G12D mutation in the stage I-III subgroup versus those in the stage IV subgroup (**Fig. 5C).**

**Fig. 5.**
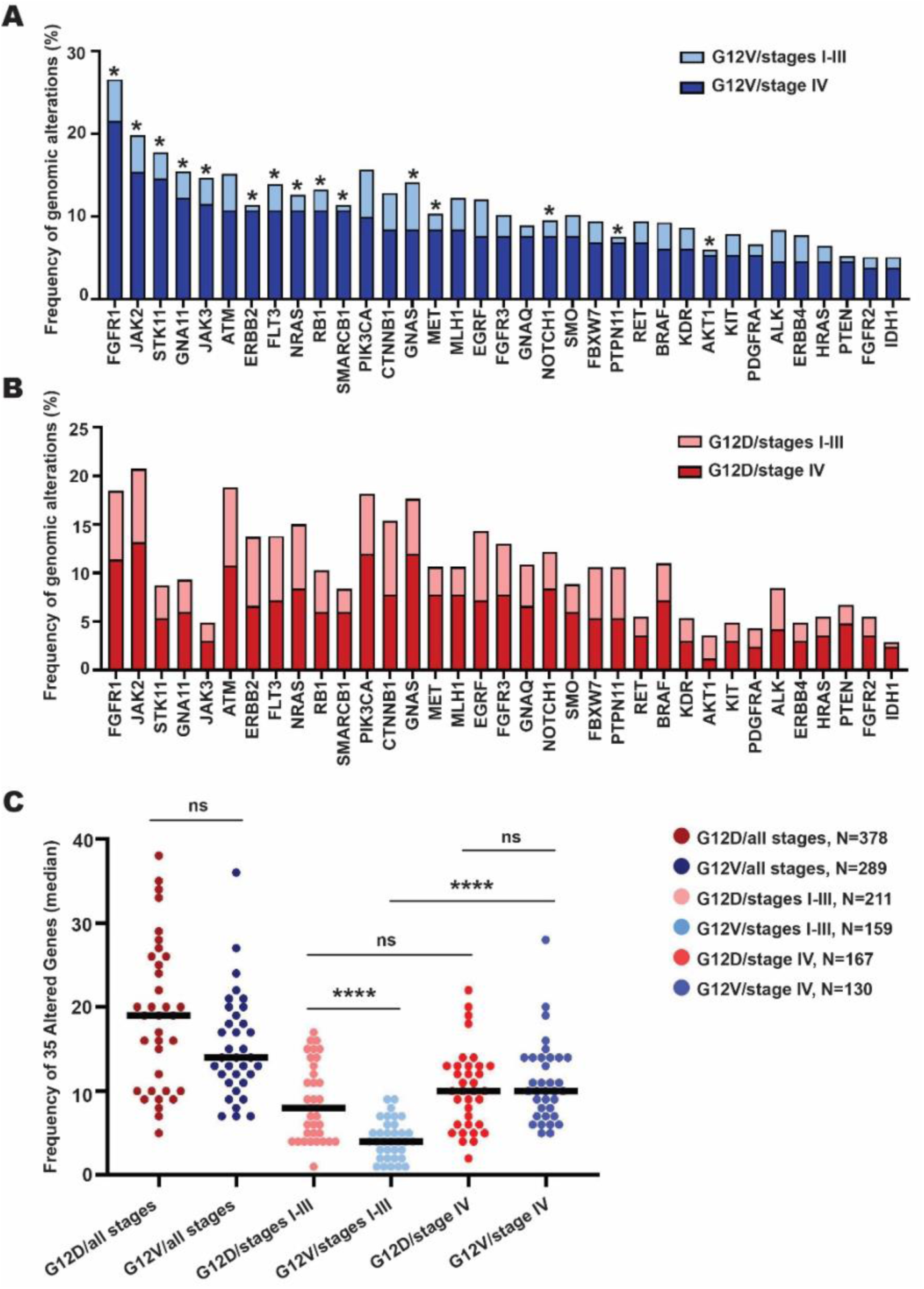
Frequency of ganomic alterations in non-driver genes in *KRAS* G12D and G12V mutant patients. Stacked bar charts displaying the frequency of genomic alterations including copy number variations or mutations of the 35 genes in GENIE cohort (excluding most commonly altered ones including *TP53*, *CDKN2A*, and *SMAD4*) for patients with **(A)** *KRAS* G12V mutation in stages I-III and IV, and **(B)** *KRAS* G12D mutation in stages I-III and IV. **(C)** Frequency of overall genomic alterations between *KRAS* G12V and G12D across stages. Each dot corresponds to the frequency of each gene in selected group and horizontal lines regarded as medians. Wilcoxon rank-sum test was used to compare between groups. False discovery rate (FDR) correction was applied for multiple comparisons, with a significance threshold of 0.05. ns = non-significant; *, *P* < 0.05; ****, *P* < 0.0001.

Next, we analyzed the overall genomic alterations across these 35 non-driver genes in the *KRAS* G12D and G12V subgroups by calculating the overall genomic alterations frequencies per group (**Fig. 5C**). Independent of stage, there were no statistically significant differences in the genomic alteration frequency of non-driver genes between patients whose tumors harbored *KRAS* G12D or G12V mutations. We asked whether the frequency of alterations in these non-driver genes was different when stratified by stage at diagnosis. Intriguingly, patients with *KRAS* G12V mutations diagnosed with localized disease had the lowest overall genomic alterations, whereas those with *KRAS* G12V diagnosed at advanced-stage had the highest number of genomic alterations (*P* < 0.001). In contrast, patients with *KRAS* G12D mutation diagnosed with stages I-III versus IV disease did not demonstrate significant differences in non-driver genomic alterations (*P* = 0.1). Thus, these results suggest that the accumulation of genomic alterations in patients with *KRAS* G12V mutations diagnosed at a later disease stage may explain the paradoxical impact of *KRAS* G12V mutation across localized and advanced stages, and might have contributed to the poor prognosis of *KRAS* G12V patients diagnosed with stage IV disease.

### Other genomic predictor of survival

To further elucidate the genomic predictors of outcomes in PDAC, we performed univariable and multivariable analyses to assess the associations between genomic factors and OS based on stage at diagnosis **(Tables 3 and 4)**. In patients diagnosed at stage I-III, univariable Cox regression analyses demonstrated that OS was significantly associated with *KRAS* G12D mutation (vs. non-G12D, P < 0.001), *KRAS* G12V mutation (vs. non-G12V, *P* = 0.02), *TP53* alterations (vs. wild-type TP53, *P* = 0.009), and *CDKN2A* alterations (vs. wild-type CDKN2A, *P* < 0.001). However, no significant associations were found for *KRAS* G12R (vs. non-G12R, *P* = 0.9), *KRAS* Q61 (vs. non-Q61, *P* = 0.5), other *KRAS* mutations (vs. non-other, P = 0.81), or *SMAD4* alterations (vs. intact SMAD4, *P* = 0.3). In multivariable Cox analysis of patients diagnosed at stages I-III, *KRAS* G12D mutation (*P* < 0.001), *KRAS* G12V mutation (*P* = 0.02), and *CDKN2A* alterations (*P* < 0.001) remained significant for OS after adjusting for other covariates **(Table 3).** In patients diagnosed at stage IV, univariable analysis found that *KRAS* G12V mutation (*P* < 0.001), *TP53* alterations (*P* = 0.003), and *CDKN2A* alterations (*P* = 0.01) were significant predictors of OS. In multivariable analysis, all three genomic alterations remain significantly associated with worse survival outcomes (*P* < 0.001, and *P* = 0.01, and *P* = 0.03, respectively) **(Table 4)**. These findings suggest that in patient diagnosed with localized PDAC, *KRAS* G12D and altered *CDKN2A* may serve as predictive biomarkers for reduced OS, while *KRAS* G12V may indicate a more favorable prognosis. In contrast, in patients diagnosed with metastatic disease, *KRAS* G12V and alterations in *TP53* and *CDKN2A* appear to confer the worst survival outcomes.

**Table 3.** Univariable and multivariable overall survival analyses of patients in stages I-III.

|  | *N* | Median survival (months) | Univariable |  | Multivariable |  |
| --- | --- | --- | --- | --- | --- | --- |
| Variables |  |  | HR (95% CI) | *P* value | HR (95% CI) | <sup>a</sup>*P* value |
| ***KRAS* mutations** |  |  |  |  |  |  |
| **G12D** | 214 | 26.7 | 1.39 (1.14-1.17) | **<0.001** | 1.99 (1.62-2.45) | **<0.001** |
| **Non-G12D** | 351 | 34.3 | - |  | - |  |
| **G12V** | 172 | 36.8 | 0.78 (0.64-0.96) | **0.02** | 0.76 (0.61-0.95) | **0.02** |
| **Non-G12V** | 393 | 28.4 | - |  | - |  |
| **G12R** | 85 | 28.4 | 1.00 (0.76-1.30) | 0.9 |  |  |
| **Non-G12R** | 480 | 31 | - |  |  |  |
| **Q61** | 32 | 33.6 | 0.87 (0.58-1.31) | 0.5 |  |  |
| **Non-Q61** | 533 | 30.6 | - |  |  |  |
| **Other *KRAS*** | 18 | 29.5 | 1.07 (0.59-1.93) | 0.81 |  |  |
| **No other *KRAS*** | 547 | 31 | - |  |  |  |
| ***TP53*** |  |  |  |  |  |  |
| **Altered** | 402 | 28.6 | 1.36 (1.09-1.69) | **0.009** | 1.23 (0.97-1.57) | 0.08 |
| **Intact** | 131 | 36 | - |  | - |  |
| ***CDKN2A*** |  |  |  |  |  |  |
| **Altered** | 172 | 23.8 | 1.61 (1.28-2.03) | **<0.001** | 1.64 (1.31-2.05) | **<0.001** |
| **Intact** | 361 | 34 | - |  | - |  |
| ***SMAD4*** |  |  |  |  |  |  |
| **Altered** | 183 | 28.6 | 1.09 (0.88-1.35) | 0.3 |  |  |
| **Intact** | 350 | 32 | - |  |  |  |

<sup>a</sup>Cox proportional hazards regression model adjusted for age, sex, race, center, and liver and thoracic metastases.

**Table 4.** Univariable and multivariable overall survival analyses of patients in stage IV.

|  | *N* | Median survival (months) | Univariable |  | Multivariable |  |
| --- | --- | --- | --- | --- | --- | --- |
| Variables |  |  | HR (95% CI) | *P* value | HR (95% CI) | <sup>a</sup>*P* value |
| ***KRAS* mutations** |  |  |  |  |  |  |
| **G12D** | 176 | 12.46 | 0.89 (0.73-1.09) | 0.2 |  |  |
| **Non-G12D** | 291 | 12.23 | - |  |  |  |
| **G12V** | 133 | 10.49 | 1.43 (1.13-1.80) | **0.001\*** | 1.44 (1.16-1.79) | **<0.001\*** |
| **Non-G12V** | 334 | 13.55 | - |  | - |  |
| **G12R** | 64 | 15.29 | 0.81 (0.62-1.07) | 0.1 |  |  |
| **Non-G12R** | 403 | 11.97 | - |  |  |  |
| **Q61** | 42 | 13.18 | 1.07 (0.76-1.52) | 0.6 |  |  |
| **Non-Q61** | 425 | 12.23 | - |  |  |  |
| **Other *KRAS*** | 13 | 11.11 | 1.01 (0.56-1.80) | 0.9 |  |  |
| **No other *KRAS*** | 454 | 12.39 | - |  |  |  |
| ***TP53*** |  |  |  |  |  |  |
| **Altered** | 357 | 11.11 | 1.47 (1.16-1.86) | **0.003\*** | 1.40 (1.06-1.84) | **0.01\*** |
| **Intact** | 85 | 16.47 | - |  | - |  |
| ***CDKN2A*** |  |  |  |  |  |  |
| **Altered** | 230 | 10.72 | 1.27 (1.04-1.56) | **0.01\*** | 1.10 (0.88-1.37) | **0.03\*** |
| **Intact** | 212 | 14.37 | - |  | - |  |
| ***SMAD4*** |  |  |  |  |  |  |
| **Altered** | 196 | 11.31 | 1.17 (0.95-1.43) | 0.1 |  |  |
| **Intact** | 246 | 13.55 | - |  |  |  |

<sup>a</sup>Cox proportional hazards regression model adjusted for age, sex, race, center, and liver and thoracic metastases.

## Discussion

While the importance of mutant *KRAS* on PDAC tumorigenesis, outcomes, and more recently, treatment options, is indisputable, the impact of codon-specific *KRAS* mutations has to date been difficult to ascertain due to a lack of datasets coupling richly annotated genomic and clinical data. In this study, we utilized data from AACR’s Project GENIE to analyze 1,032 patients with PDAC and specifically test how codon-specific *KRAS* mutations are associated with co-occurring genomic alterations and PDAC outcomes across disease stages. We found that the *KRAS* G12V mutation is associated with significantly longer survival in patients diagnosed with localized PDAC, yet is associated with shorter survival in patients with metastatic disease at the time of diagnosis. Multivariable analyses confirm the paradoxical impact of the *KRAS* G12V mutation across different disease stages, regardless of metastatic patterns, use of different treatment modalities, or alterations in the common tumor suppressors *TP53*, *CDKN2A*, and *SMAD4*. Detailed analyses of additional genomic alterations indicate that changes in frequencies of ’non-driver’ genes may explain the paradoxical impact of the *G12V* mutation on patient outcomes in both localized and advanced PDAC. Taken together, our findings highlight the importance of considering codon-specific *KRAS* mutations as key factors in PDAC prognosis based on stage at diagnosis.

To date, studies evaluating the impact of codon-specific *KRAS* mutations in PDAC have produced conflicting results, in part due to the fact that these studies often evaluated patient populations that different based on stage at diagnosis or lacked important clinical annotations (6,7,14–16,19). More recently, studies have emerged that provide detailed depictions of more uniform patient populations. Consistent with our findings, several studies evaluating patients with localized disease have now reported that *KRAS* G12D mutations are associated with significantly worse outcomes compared to non-G12D mutations (6,14,15). Most recently, McIntyre et al. compared overall survival in 397 PDAC patients diagnosed at stage I-III, and found that those diagnosed at stage I with *KRAS* G12D mutations had significantly worse survival compared to patients with G12R and G12V mutations (16). Our results largely align with this study, showing that patients with *KRAS* G12V mutation have the best survival outcomes and patients with G12D mutation the worst in patients diagnosed with localized disease. Unlike their study, we found similar survival rates for *KRAS* G12D and G12R mutations. This discrepancy may be due to the limited patient numbers in their cohort and their focus on stage I patients, which had a higher frequency of *KRAS* G12R mutation.

In contrast, in patients diagnosed with advanced metastatic disease, we found that *KRAS* G12V was associated with worse overall survival, which conflicts with several previous reports. One study by Pan et al. (893 patients), which focused on patients with stage IV PDAC, highlighted the adverse impact of the Q61 mutations on survival (19). Another cohort of 578 patients that included those diagnosed with both localized and metastatic disease observed that the *KRAS* Q61 mutations were associated with shorter OS across all stages of PDAC, while *KRAS* G12D was linked to poorer OS in stage IV disease (7). In alignment with our study, Cheng et al. reported that the *KRAS* G12V mutation was associated with poor prognosis in advanced pancreatic cancer (210 patients) (24). Diehl et al. also reported longer OS in metastatic PDAC patients with the G12R mutation compared to non-G12R patients (25). However, our univariable analysis did not identify the *KRAS* G12R mutation as an independent prognostic variable in stage IV disease. In summary, our findings indicate that the *KRAS* G12V mutation is associated with a poor prognosis in stage IV disease, which contradicts several studies. This discrepancy may arise from the patient cohort being sourced from a single institute or due to the limited number of patients in their cohorts.

Over the last decade, growing research has elucidated that each *KRAS* mutation has distinct biochemical features that affect GTP hydrolysis, nucleotide exchange, and effector interactions, resulting in varied neoplastic phenotypes (26–30). For instance, both valine (G12V) and aspartic acid (G12D) substitutions at codon 12 decrease the affinity of *KRAS* for the effector RAF. However, the G12V mutation is observed to cause a more substantial reduction of GTPase activity through both intrinsic hydrolysis and GAP-mediated hydrolysis (26). Despite these insights, evidence on the precise distinctions between the nature of *KRAS* G12D and G12V mutations and the factors influencing them remains scarce, making this paper the first to highlight the clinical contrast between *KRAS* G12D and *KRAS* G12V in PDAC.

The rapid development and adoption of targeted *KRAS* inhibition through various approaches, including G12C inhibitors (31,32), G12D inhibitor (33), pan-*KRAS* inhibitor (34), pan-RAS inhibitors (35,36), and *KRAS* degraders (37) will almost certainly alter how PDAC is treated across stages. At present, the impact of different *KRAS* mutation subtypes on PDAC prognosis and their resultant therapeutic vulnerabilities has remains uncertain, representing a critical knowledge gap in the interpretation and design of ongoing and emerging clinical trials. Our findings depict distinct clinical implications of codon-specific *KRAS* mutations in localized and advanced stages of pancreatic cancer, thus suggesting codon-specific *KRAS* mutation status as a potential independent prognostic factor of therapeutic response and progression-free survival with RAS inhibitors and other treatment regimens. Our study thus contributes to an overall improved understanding of the impact of these codon-specific mutations, which will be crucial to ongoing and future studies testing *KRAS* inhibition in PDAC across disease stages.

Notably, the frequency of genomic alterations reported in this study for the common driver genes (*KRAS*, *TP53*, *CDKN2A*, and *SMAD4*) is consistent with other large datasets (14,19,25,38). Similar to previous studies (16,19,39), alterations in each of these tumor suppressor genes were individually associated with worse survival outcomes. Furthermore, *KRAS* mutations, when coupled with genetic alterations in *TP53*, *SMAD4*, and *CDKN2A*, have been shown to accelerate PDAC progression and worsen survival outcomes (15,19,40). In consistence with Pan et al, we also observed that *CDKN2A* alterations alone, or in combination with *SMAD4* and *TP53*, in *KRAS* mutant patients were associated with the worst survival outcomes in PDAC prognosis (19). These consistencies support the conclusions of this study, which we suspect are only possible to fully elucidate in a large, diverse dataset that couples richly annotated genomic and clinical data.

While we did not find any association between the codon-specific *KRAS* mutations and alterations in *TP53*, *CDKN2A*, and *SMAD4* as the main driver genes in PDAC, we interestingly uncovered an association between the prognostic impact of *KRAS* G12V in localized and advanced disease stages and differential genomic alterations, primarily driven by CNVs or mutations in 35 non-driver genes in PDAC. One potential interpretation of this finding is that while *KRAS* G12V dictates a better prognostic impact on PDAC patient diagnosed with localized disease, this advantage disappeared when accompanied by higher genomic alterations in other non-driver genes in advanced disease. However, future studies are required to find any association between codon-specific *KRAS* mutations and whole genomic alterations in PDAC.

Our study’s strengths include a large patient cohort from multiple institutions and a comprehensive analysis of diverse clinical variables across all disease stages. However, despite the large sample size, limitations remain. First, we were unable to address the role of rare codon-specific *KRAS* mutations, such as *KRAS* Q61H, Q61R, and Q61L, in subgroup analyses due to the limited number of patients with these mutations. Second, we focused on genomic alterations of only 35 non-driver genes. These genes were selected based on the fact that all were included within the panels used by the four centers from which data were derived, and thus were not biased by missing data. However, further studies are needed to assess global genomic alterations in PDAC patient samples in the context of codon-specific *KRAS* mutations and prognosis. Third, it is a retrospective study and detailed imaging and surgical data, and dynamic variables such as smoking status were not captured. Finally, the heterogeneity of treatment data restricts us to assess the impact of codon-specific KRAS mutations on different chemotherapy, radiation, and investigational treatment responses. Despite these limitations, this study represents the largest effort to evaluate codon-specific *KRAS* mutations across stages of PDAC, leveraging robust clinical annotations to help adjust for confounding.

In summary, our analysis of the AACR’s GENIE Biopharma Consortium Pancreas v1.2 dataset with comprehensive genomic and clinical data revealed that codon-specific *KRAS* mutations impact PDAC outcomes differently based on disease stage at diagnosis. As research on RAS inhibitors progresses, these findings provide valuable contextual insights into survival outcomes associated with codon-specific *KRAS* mutations based on existing therapeutic approaches.

## Authors’ Contributions

**S. Raji**: Conceptualization, designed the study, methodology, data analysis and interpretation, visualization, writing original and final manuscript. **H. Zaribafzadeh**: Data curation and management, software, data analysis and interpretation, methodology, visualization, writing and editing. **T. Jones**: data analysis and interpretation, methodology. **E. Kanu**: Data curation, writing and editing. **K. Tong**: Data curation, analysis and interpretation. **A. Fletcher**: Project administration, writing and editing. **T. C. Howell**: Data curation and analysis. **S.J. McCall**: Resources, project administration. **J.R. Marks**: Data analysis and interpretation, review and editing manuscript. **B. Rogers**: Data analysis and interpretation. **D. Niedzwiecki**: Supervision, statistical analysis and validation, review and editing. **P.J. Allen**: Conceptualization, supervision, data analysis and interpretation. **D.P. Nussbaum**: Conceptualization, designed the study, methodology, data analysis and interpretation, supervision, writing, reviewing and editing manuscript. **Z. Kabiri**: Conceptualization, designed the study, methodology, supervision, data analysis and interpretation, project administration, writing original and final manuscript.

## Acknowledgments

The authors express their gratitude to the AACR for its financial and material support in establishing the AACR Project GENIE registry and to the members of the AACR Project GENIE consortium for their dedication to data sharing. Also they acknowledge the valuable contributions of the GENIE Coordinating Center, Sage Bionetworks, and/or cBioPortal staff in data development. Zahra Kabiri was supported by the Duke Surgery Department start-up fund and National Cancer Institute (K22CA266753) grant.

