## Supplementary Fig. S1 for "Prognostic Implications of Codon-Specific *KRAS* Mutations in Localized and Advanced Stages of Pancreatic Cancer"

**Supplementary Fig. S1. Study design.** The flowchart shows GENIE cohort patient selection criteria in our study.

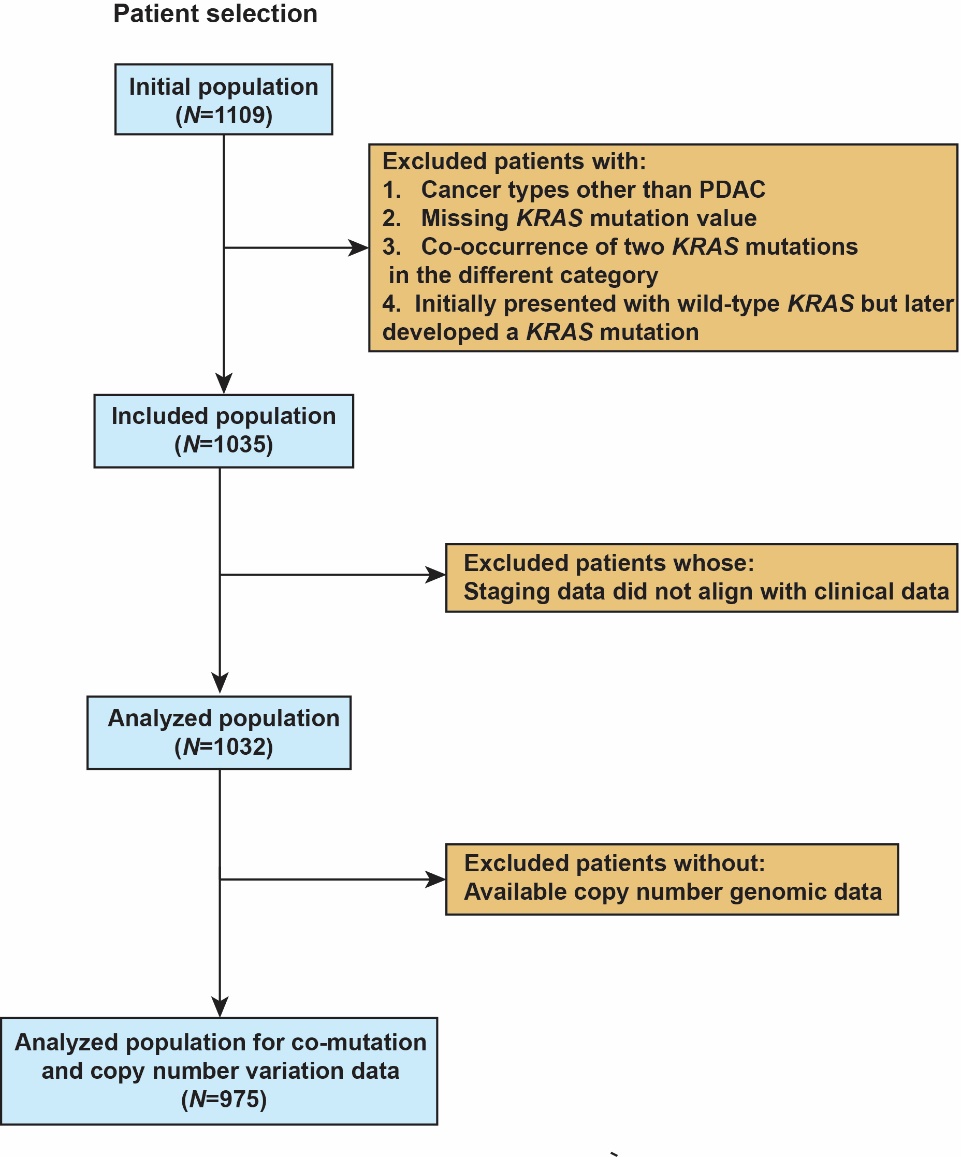
