## Supplementary Fig. S2 for "Prognostic Implications of Codon-Specific *KRAS* Mutations in Localized and Advanced Stages of Pancreatic Cancer"

**Supplementary Fig. S2.** **Kaplan–Meier curves for overall survival**. Kaplan-Meier survival analyses of patients with **(A)** mutated *KRAS* versus wild-type *KRAS* (HR, 1.26; 95% CI, 1.005-1.59), **(B)** altered *TP53* versus intact *TP53* (HR, 1.44; 95% CI, 1.23-1.69), **(C)** altered *CDKN2A* versus intact *CDKN2A* (HR, 1.70; 95% CI, 1.46-1.98), **(D)** altered *SMAD4* versus intact *SMAD4* (HR, 1.22; 95% CI, 1.05-1.41). **(E)** Kaplan-Meier survival analyses of tumor suppressor alterations in patients with mutant *KRAS*. PDAC, pancreatic ductal adenocarcinoma; Intact genes pointed to samples without copy number, structural variant, or single-nucleotide variant events detected; Altered, indicates copy-number losses or mutation of gene; HR, hazard ratio; *, P < 0.05 was considered statistically significant, calculated by log-rank test.

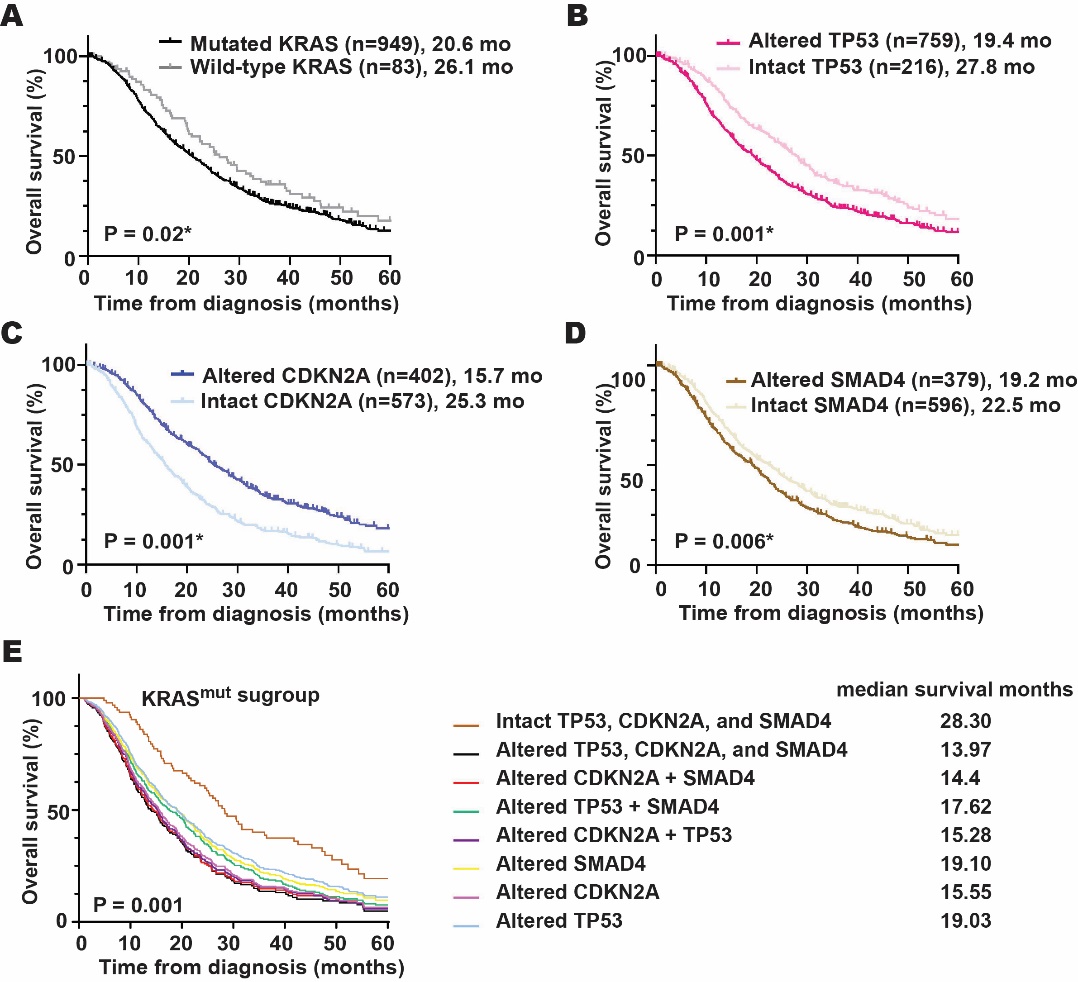
