## Supplementary Fig. S3 for "Prognostic Implications of Codon-Specific *KRAS* Mutations in Localized and Advanced Stages of Pancreatic Cancer"

**Su****pplementary Fig. S3. Forest plot of multivariable Cox proportional hazards models of overall survival within subgroups.** **(A)** The HR for OS comparing patients with *KRAS* G12V versus G12D was estimated using a multivariable Cox model across the entire cohort based on PDAC stage groups (I-III and IV separately), including analyses of subgroups with alterations in *TP53*, *CDKN2A*, and *SMAD4*; liver metastasis; patients who did not undergo surgery; those who received or did not receive adjuvant or neoadjuvant chemotherapy in stages I-III, and those who received any first-line treatment in stage IV. We included variables with a *P* value < 0.05 in the univariable analysis adjusting for age, sex, race, and center. OS, overall survival; HR, hazard ratio; CI, confidence interval; Adj, adjuvant; neoadj, neoadjuvant.

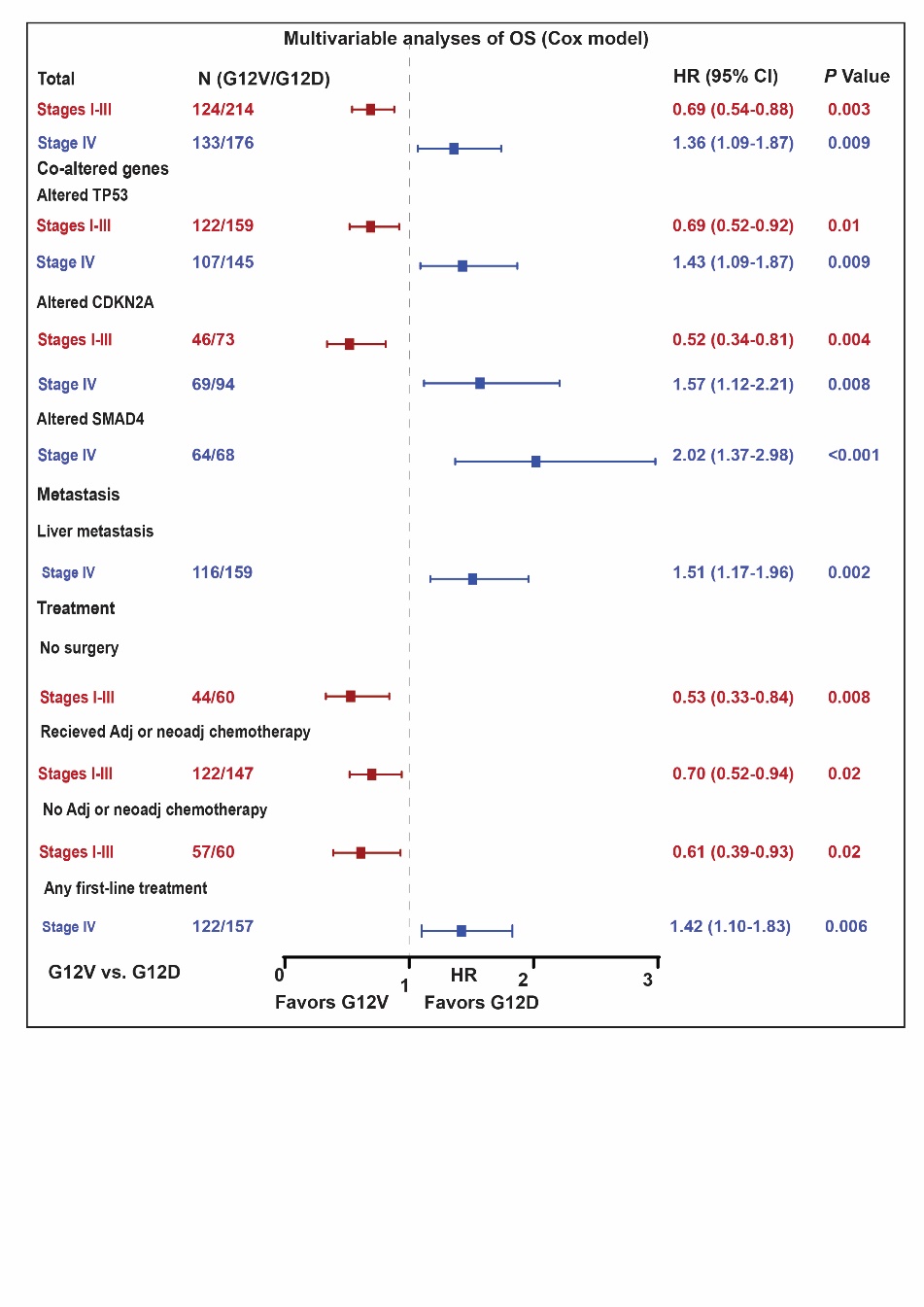
