## Supplementary Table S1 for "Prognostic Implications of Codon-Specific *KRAS* Mutations in Localized and Advanced Stages of Pancreatic Cancer"

**Supplementary Table S1. Gene panels.** Summary of gene panels used by institution.

| Gene panel | *N* (%)  *N*=1032 |
| --- | --- |
| DFCI |  |
| DFCI-ONCOPANEL-1 | 18 (2) |
| DFCI-ONCOPANEL-2 | 118 (11) |
| DFCI-ONCOPANEL-3 | 177 (17) |
| DFCI-ONCOPANEL-3.1 | 99 (10) |
| MSKCC |  |
| MSK-IMPACT341 | 4 (<1) |
| MSK-IMPACT410 | 167 (16) |
| MSK-IMPACT468 | 329 (32) |
| UHN |  |
| UHN-48-V1 | 37 (4) |
| UHN-555-PAN-GI-V1 | 5 (<1) |
| UHN-555-V1 | 12 (1) |
| UHN-OCA-V3 | 3 (<1) |
| VICC |  |
| VICC-01-T5A | 8 (1) |
| VICC-01-T7 | 55 (5) |

DFCI, Dana Farber Cancer Institute; MSKCC, Memorial Sloan Kettering Cancer Center; UHN, Princess Margaret Cancer Centre-University Health Network; VICC, Vanderbilt Ingram Cancer Center.
