## Supplementary Table S2 for "Prognostic Implications of Codon-Specific *KRAS* Mutations in Localized and Advanced Stages of Pancreatic Cancer"

**Supplementary Table S2. KRAS mutations**. The detailed list of KRAS mutations frequency in GENIE dataset (N=951).

| *KRAS* mutations | *N* (%)  *N*=951* |
| --- | --- |
| G12D | 390 (41) |
| G12V | 305 (32) |
| G12R | 149 (16) |
| Q61H | 42 (4.4) |
| Q61R | 20 (2.1) |
| G12C | 12 (1.4) |
| Q61L | 8 (0.8) |
| G13D | 8 (0.8) |
| Q61K | 4 (0.4) |
| G12A | 3 (0.3) |
| G12I | 2 (0.2) |
| D33H | 1 (0.1) |
| G13V | 1 (0.1) |
| G13E | 1 (0.1) |
| G12L | 1 (0.1) |
| A11G | 1 (0.1) |
| A146V | 1 (0.1) |
| G12F | 1 (0.1) |
| L52_L53insWIFSTQ | 1 (0.1) |

* Two patients had co-occurrence of *KRAS* mutations in same “other” category.
