## Supplementary Table S3 for "Prognostic Implications of Codon-Specific *KRAS* Mutations in Localized and Advanced Stages of Pancreatic Cancer"

| All stages | Mutant *KRAS* cohort | | | |
| --- | --- | --- | --- | --- |
| Tumor suppressor alterations | *N* | Median survival months | HR (CI 95%) | **P* value |
| Absence of alterations in *TP53*, *CDKN2A*, and *SMAD4* (reference) | 95 | 28.30 | - | - |
| Alterations in *TP53*, *CDKN2A*, and *SMAD4* | 189 | 13.97 | 2.09 (1.62-2.70) | **<0.0001** |
| Altered *CDKN2A* + *SMAD4* | 213 | 14.40 | 2.01 (1.57-2.57) | **<0.0001** |
| Altered *TP53* + *SMAD4* | 299 | 17.62 | 1.72 (1.37-2.18) | **<0.0001** |
| Altered *CDKN2A* + *TP53* | 332 | 15.28 | 1.97 (1.58-2.46) | **<0.0001** |
| Altered *SMAD4* | 358 | 19.10 | 1.56 (1.24-1.97) | **0.0006** |
| Altered *CDKN2A* | 380 | 15.55 | 1.91 (1.54-2.37) | **<0.0001** |
| Altered *TP53* | 727 | 19.03 | 1.50 (1.21 – 1.86) | **0.001** |

**Supplementary Table S3.** Univariable analysis of OS by combination of tumor suppressor alterations in patients with mutant KRAS across all stages.

**P* values by log-rank test.
