## Supplementary Table S4 for "Prognostic Implications of Codon-Specific *KRAS* Mutations in Localized and Advanced Stages of Pancreatic Cancer"

**Supplementary Table S4.** Frequencies of genomic alterations and in codon-specific KRAS mutations in all stages.

| Variables | G12D  (*N*=390) | G12V  (*N*=305) | G12R  (*N*=149) | Q61  (*N*=74) | Other  (*N*=73) | ^*^*P* value |
| --- | --- | --- | --- | --- | --- | --- |
| Altered *TP53* | 304 (80.4%) | 229 (79.2%) | 107 (78.7%) | 62 (86.1%) | 25 (83.3%) | 0.7 |
| Unknown | 12 | 16 | 13 | 2 | 1 |  |
| Altered *CDKN2A* | 167 (44.2%) | 115 (39.8%) | 57 (41.9%) | 30 (41.7%) | 11 (36.7%) | 0.8 |
| Unknown | 12 | 16 | 13 | 2 | 1 |  |
| Altered *SMAD4* | 141 (37.3%) | 113 (39.1%) | 64 (47.1%) | 25 (34.7%) | 15  (50%) | 0.2 |
| Unknown | 12 | 16 | 13 | 2 | 1 |  |

^*^ Pearson Chi-Square Test (χ²)
