## Supplementary Table S5 for "Prognostic Implications of Codon-Specific *KRAS* Mutations in Localized and Advanced Stages of Pancreatic Cancer"

**Supplementary Table S5**. Frequencies of genomic alterations and in codon-specific KRAS mutations in patients diagnosed in stages I-III.

| Variables | G12D  (*N*=214) | G12V  (*N*=172) | G12R  (*N*=85) | Q61  (*N*=32) | Other  (*N*=19) | ^*^*P* value |
| --- | --- | --- | --- | --- | --- | --- |
| Altered *TP53* | 159 (75.4%) | 122 (76.7%) | 63 (80.8%) | 26 (86.7%) | 14 (77.8%) | 0.6 |
| Unknown | 3 | 13 | 7 | 2 | 0 |  |
| Altered *CDKN2A* | 73 (34.6%) | 46 (28.9%) | 31 (39.7%) | 10 (33.3%) | 4  (22.2%) | 0.4 |
| Unknown | 3 | 13 | 7 | 2 | 0 |  |
| Altered *SMAD4* | 73 (34.6%) | 49 (30.8%) | 37 (47.4%) | 8 (26.7%) | 8  (44.4%) | 0.08 |
| Unknown | 3 | 13 | 7 | 2 | 0 |  |
