## Supplementary Table S6 for "Prognostic Implications of Codon-Specific *KRAS* Mutations in Localized and Advanced Stages of Pancreatic Cancer"

**Supplementary Table S6**. Frequencies of genomic alterations and in codon-specific KRAS mutations in patients diagnosed in stage IV.

| Variables | G12D  (*N*=176) | G12V  (*N*=133) | G12R  (*N*=64) | Q61  (*N*=42) | Other  (*N*=14) | ^*^*P* value |
| --- | --- | --- | --- | --- | --- | --- |
| Altered *TP53* | 145 (76.8%) | 107  (82.3%) | 44  (75.9%) | 36  (85.7%) | 11  (91.7%) | 0.3 |
| Unknown | 9 | 3 | 6 | 0 | 1 |  |
| Altered *CDKN2A* | 94  (56.3%) | 69  (53.1%) | 26  (44.8%) | 20 (47.6%) | 8  (58.3%) | 0.5 |
| Unknown | 9 | 3 | 6 | 0 | 1 |  |
| Altered *SMAD4* | 68  (40.7%) | 64  (49.2%) | 27 (46.6%) | 17 (40.5%) | 7  (58.3%) | 0.4 |
| Unknown | 9 | 3 | 6 | 0 | 1 |  |
