## Supplementary Table S7 for "Prognostic Implications of Codon-Specific *KRAS* Mutations in Localized and Advanced Stages of Pancreatic Cancer"

**Supplementary Table S7.** Adjuvant chemotherapy treatments in stages I-III.

| Regimen/Drugs | Overall (*N*=233) | G12D (*N*=131) | G12V (*N*=102) |
| --- | --- | --- | --- |
| 5FU/gem/cape | 169 (72.5%) | 88 (67.2%) | 81 (79.4%) |
| FOLFIRINOX | 28 (12.0%) | 17 (13.0%) | 11 (10.8%) |
| Gem/abraxane | 18 (7.7%) | 14 (10.7%) | 4 (3.9%) |
| Cis-based | 3 (1.3%) | 1 (0.8%) | 2 (2.0%) |
| FOLFOX/GEMOX/CAPOX | 7 (3.0%) | 4 (3.1%) | 3 (2.9%) |
| Investigational Drug | 6 (2.6%) | 6 (4.6%) | 0 (0%) |
| Immunotherapy+ | 1 (0.4%) | 0 (0%) | 1 (1.0%) |

5FU/gem/cape, 5FU (Fluorouracil)/Gemcitabine/Capecitabin; Cis-based, cisplatin-based; FOLFOX (fluorouracil, leucovorin calcium, and oxaliplatin)/GEMOX (gemcitabine and oxaliplatin)/CAPOX (capecitabine and oxaliplatin).
