## Supplementary Table S8 for "Prognostic Implications of Codon-Specific *KRAS* Mutations in Localized and Advanced Stages of Pancreatic Cancer"

**Supplementary Table S8**. Neoadjuvant chemotherapy treatments in stages I-III.

| Regimen/Drugs | Overall (*N*=279) | G12D (*N=*157) | G12V (*N*=122) |
| --- | --- | --- | --- |
| 5FU/gem/cape | 9 (3.2%) | 3 (1.9%) | 6 (4.9%) |
| Cis-based | 2 (0.7%) | 1 (0.6%) | 1 (0.8%) |
| FOLFIRINOX | 156 (55.9%) | 95 (60.5%) | 61 (50.0%) |
| FOLFOX/GEMOX/CAPOX | 8 (2.9%) | 4 (2.5%) | 4 (3.3%) |
| Gem/abraxane | 68 (24.4%) | 34 (21.7%) | 34 (27.9%) |
| Investigational Drug | 32 (11.5%) | 18 (11.5%) | 14 (11.5%) |
| Other | 4 (1.4%) | 2 (1.3%) | 2 (1.6%) |

5FU/gem/cape, 5FU (Fluorouracil)/Gemcitabine/Capecitabin; Cis-based, cisplatin-based; FOLFOX (fluorouracil, leucovorin calcium, and oxaliplatin)/GEMOX (gemcitabine and oxaliplatin)/CAPOX (capecitabine and oxaliplatin).
