## Supplementary Table S9 for "Prognostic Implications of Codon-Specific *KRAS* Mutations in Localized and Advanced Stages of Pancreatic Cancer"

**Supplementary Table S9**. Most common first-line treatments in stage IV.

| Regimen/Drugs | Overall (*N*=77) | G12D (*N*=39) | G12V (*N*=38) |
| --- | --- | --- | --- |
| FOLFIRINOX | 58 (75.3%) | 30 (76.9%) | 28 (73.7%) |
| Gem/abraxane | 12 (15.6%) | 6 (15.4%) | 6 (15.8%) |
| Investigational Drug | 3 (3.9%) | 0 (0%) | 3 (7.9%) |
| 5FU/gem/cape | 2 (2.6%) | 2 (5.1%) | 0 (0%) |
| Other | 2 (2.6%) | 1 (2.6%) | 1 (2.6%) |

5FU/gem/cape, 5FU (Fluorouracil)/Gemcitabine/Capecitabine.
